# Photobiomodulation promotes wound healing and functional improvement following lumbar decompression surgery: a double-blinded, placebo-controlled study

**DOI:** 10.64898/2026.07.15.26357882

**Authors:** Joshua Rivera, Yan Zhou, Lara Sak, Fiona Pudewa, Jihoon Lee, Mark T. Yamamoto, Habin Yoo, Meachelle Lum, Michelle Zhang, Aakash Patel, Louise E. Vandenberghe, Sara K. Fenn, Yuanchen Wang, Brooke Bailey, Sandra M. Holley, Andrew C. Vivas, Langston T. Holly, Daniel C. Lu

**Affiliations:** David Geffen School of Medicine at UCLA, Los Angeles, CA, USA; School of Medicine, University of Colorado Anschutz Medical Campus, Aurora, CO, USA; Department of Medicine, California University of Science and Medicine, Los Angeles, CA, USA; Department of Neurosurgery, University of California, Los Angeles, Los Angeles, CA, USA; University of California, Los Angeles, Los Angeles, CA, USA; School of Medicine, University of Pittsburgh, Pittsburgh, PA, USA

**Keywords:** photobiomodulation, lower back pain, wound healing, leg pain, scar, lumbar decompression

## Abstract

**Objective:** Photobiomodulation therapy has emerged as a promising modality to facilitate scar healing and pain management in dermatology and plastic surgery. However, its role in postoperative care following spine surgeries remains understudied. This double-blinded, placebo-controlled study aimed to investigate the effects of photobiomodulation in patients with chronic lower back pain undergoing lumbar decompression, with postoperative wound healing as the primary outcome and pain reduction and functional recovery as secondary outcomes.

**Methods:** Patients were randomized to receive either active photobiomodulation braces (N=13) or placebo braces (N=12). Follow-up assessments were performed at 2, 4, 6, 8, and 12 weeks postoperatively. Outcomes included wound healing (Stony Brook Scar Evaluation Scale), back and leg pain (Visual Analog Scale), quality of life (EuroQol 5D), and functional status (Oswestry Disability Index).

**Results:** Compared to the placebo group, the photobiomodulation treatment group had a 4.12-fold cumulative improvement in final scar scores, with significant between-group differences at postoperative weeks 6, 8, and 12 (p = 0.0062, 0.010, 0.042). Among patients with severe preoperative disability, treatment resulted in a 1.89-fold faster improvement in back pain (p=0.025) and a 1.80-fold faster improvement in ODI scores (p=0.025); and superior treatment effect on wound healing were again observed at weeks 6, 8, and 12. Among patients with poor initial scars, treatment led to a significantly better scar outcome than placebo at week 6 and a 1.94-fold faster EQ5D improvement (p=0.052), with significant gains observed as early as two weeks after surgery. There were no adverse events associated with photobiomodulation treatment.

**Conclusions:** Photobiomodulation significantly promoted postoperative wound healing following lumbar decompression surgery, with therapeutic benefits preserved even in patients with poor baseline scar scores and functional impairment. This indicates that the efficacy of photobiomodulation is not limited by the initial scar condition or disability, supporting its broad clinical applicability. Additionally, patients with severe preoperative disability experienced greater benefits from photobiomodulation than placebo, including faster reduction in back pain and more rapid improvement in functional capacity, highlighting its role in postoperative pain management and rehabilitation. These therapeutic effects are likely mediated by photobiomodulation-induced reduction of inflammation and enhancement of tissue repair. Together, this study suggests that photobiomodulation can be a promising adjunct therapy to facilitate postoperative recovery in patients undergoing spine surgery.

## INTRODUCTION

Lower back pain is the leading cause of disability worldwide, impacting approximately 619 million people globally ^1, 2^. The increasing burden of lower back pain has contributed to a steady rise in the number of surgical spine operations. Despite such increasing prevalence of spine operations, effective postoperative pain management remains a challenge. While various post-operative treatment plans exist to aid in recovery, including pharmacotherapy, physiotherapy, and the use of back braces, 10-40% of patients experience persistent back pain following spine surgery ^3, 4^.

The exact underlying mechanisms contributing to chronic postoperative pain remain poorly understood but are believed to be multifactorial, including epidural fibrosis, arachnoiditis, nucleus changes, sensitization of neurons, and other psychological and structural factors ^5^. Given the substantial proportion of patients who experience persistent pain, it is imperative that we optimize patient recovery and pain management following surgery.

One emerging treatment modality to address postoperative pain and inflammation is photobiomodulation therapy ^6^. Photobiomodulation, previously referred to as low-level laser therapy, was first discovered in the 1967 by Endre Mester ^7^ and has since been used in clinical practice to aid in the treatment of various diseases including diabetes, neural disease, musculoskeletal conditions, dentistry, and dermatologic conditions ^8, 9^. The mechanism behind photobiomodulation is thought to involve activation of mitochondrial cytochrome c oxidase, which in turn modulates ATP, reactive oxygen species, and calcium, ultimately altering gene expression, growth factor release, and cytokine release ^10^.

Several studies have investigated the role of photobiomodulation in promoting wound healing and treating inflammatory pain. Using a mouse model, do Valle et al. found that photobiomodulation resulted in vascular changes that accelerated tissue repair ^11^, while Pigatto et al. demonstrated the anti-nociceptive and anti-inflammatory effects of photobiomodulation in treating acute pain ^12^. While previous studies have shown efficacy of laser therapy in managing spinal cord injury and chronic back pain ^13, 14^, none to our knowledge have investigated the use of photobiomodulation in postsurgical wound and functional recovery. In this study, we investigated the effects of photobiomodulation in patients with chronic lower back pain undergoing lumbar surgical decompression.

## METHODS

### Study Design and Intervention

This study is a double-blinded, placebo-controlled randomized clinical trial with two parallel groups (treatment vs. placebo) and a superiority framework. It was performed at one academic institution with three enrolling surgeons in the United States between August 2023 and April 2025. This manuscript reports results from patients undergoing minimally invasive lumbar decompression in the registered trial. Institutional review board (IRB) approval was obtained from the UCLA IRB (IRB#23-000444), and informed consent was obtained from all participants. The trial design followed CONSORT guideline ^15^ (**Figure 1**) and was registered at ClinicalTrials.gov (NCT06282770; https://clinicaltrials.gov/study/NCT06282770). The trial protocol and statistical analysis plan are available at ClinicalTrials.gov. The braces were purchased from Capillus, Inc. (Capillus, Miami, FL). Patients were randomized into 2 groups with one receiving a photobiomodulation Capillus Laser Therapy Lumbar Brace (Capillus, Miami, FL) with active lasers (treatment group) and the other receiving a placebo brace with reduced energy (placebo group). The brace is composed of a laser lumbar support orthosis (LLSO) with a laser module that is integrated into a removable insert (**Figure 2A**). The laser module is composed of 84 laser diodes.

**Figure 1.**
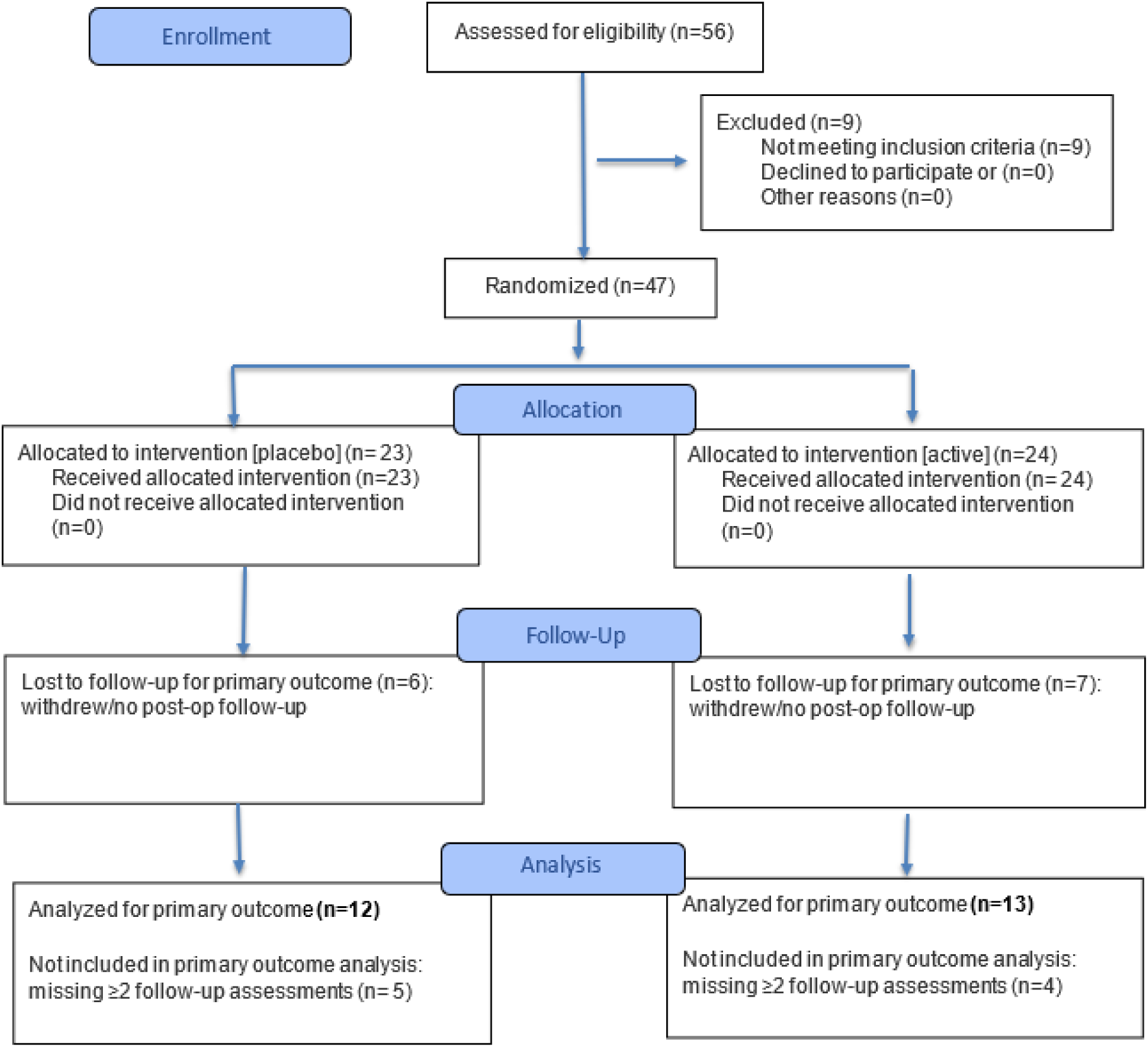
CONSORT 2025 flow diagram for patients receiving minimally invasive lumbar decompression surgery in the registered randomized clinical trial, showing the process of enrollment, allocation, follow-up, and analysis.

**Figure 2.**
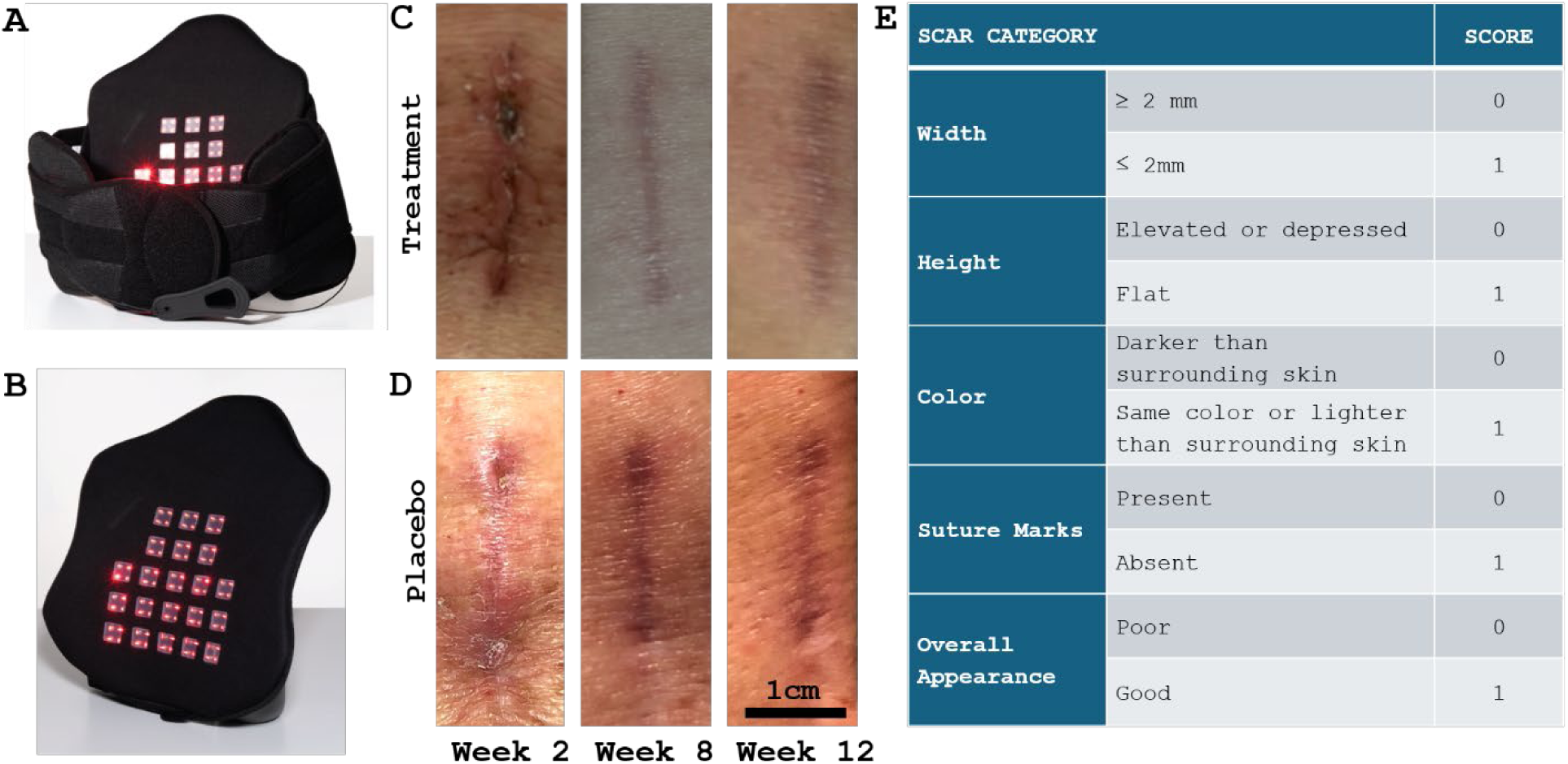
A: Image of an assembled laser brace which is composed of a lumbar support orthosis and a removable insert with laser module. **B:** Removable insert. The light-emitting module comprises 21 emitter modules, each containing four red-light diodes (84 diodes total). Each treatment diode emits low-intensity red light (4-5 mW) at 650 nm, for a total optical output of ∼420 mW. **C&D:** Representative scar images from the laser brace treatment group and placebo group. **C:** Treatment group. **D:** Placebo group. Scar images from a representative patient in each group are shown at postoperative week 2, 8, and 12 (left to right). All images are shown at the same scale for comparison. **E:** Stony Brook Scar Evaluation Scale (adapted from Singer et al., 2007).

Each diode produces low-intensity (treatment, 4-5 mW, or placebo, 0.015 mW) red light at a wavelength of 650 nm. The placebo laser diodes emitted visible red light and were visually indistinguishable from the active device. To achieve this reduction in the placebo device, additional resistors were incorporated into the laser diode driver circuit, lowering the laser output to a non-therapeutic level. This output was sufficient to emit visible red light so that the device appeared active, while avoiding delivery of any therapeutically effective dose. The laser module is powered by a rechargeable lithium-ion battery (3,500 mAh) with a preset timer of 12 minutes. Participants were instructed to wear the brace with the light on three times per day for 12 weeks, regardless of group assignment. Patients enrolled in the study were trained on use when receiving the belt and had serial check-ins with the clinic and research team at each follow-up. Participants were encouraged to ask questions throughout study duration through the use of the EMR system and provide photos with any issues. The primary outcome was postoperative wound healing and secondary outcomes were pain reduction and functional recovery.

Wound healing was assessed at baseline (preoperatively) and at 2, 4, 6, 8, and 12 weeks after surgery using Stony Brook Scar Evaluation Scale^16^ (range 0-5, higher scores indicating better healing) (**Figure 2E**). Scars were independently scored by two medical specialists using REDCap- uploaded scar photographs^17, 18^. Patient reported outcomes were collected through REDCap electronic questionnaires on the same schedule and included Visual Analog Scale (VAS) back and leg pain (0-10, 0 being no pain and 10 being worst pain), EuroQol 5D (EQ5D; 0-100, 0 being worst imaginable health state and 100 being the best state), and Oswestry Disability Index (ODI; 0- 100%, higher percentage indicating higher level of disability). Harms were defined as adverse events related to brace use and were assessed at each follow-up by clinical evaluation and patient report.

### Patient Population

Patients during the study period were consecutively enrolled, and sample size was determined based on the expected patient volume over a two-year time period in order to ensure feasibility across participating surgeons. Inclusion criteria for enrollment were ages 18+ years and undergoing minimally invasive lumbar decompression surgery at UCLA, chronic lower back pain >3 months with a self-reported pain severity >4 at screening. Exclusion criteria included tumors, infection, trauma, skin conditions precluding brace use, and severe spondylolisthesis, spondylolysis, spinal stenosis, ankylosing spondylitis. Participants who did not provide any number of post-operative follow-up data were considered lost to follow-up. Patient exclusion criteria was optimized to enhance rigor of the significance of the statistical analysis.

### Randomization and blinding

Patients were randomly assigned in a 1:1 ration to the treatment or placebo group using a computer-generated randomization algorithm (MATLAB), ensuring equal probability of group assignment. Patients were assigned sequential study codes, and the allocation key linking study codes to treatment assignments was password-protected and maintained by the principal investigator. All personnel involved in enrollment, brace distribution, outcome assessment, and data analyses remained blinded to group assignment throughout the study.

### Statistical Analysis

Statistical analyses were performed on all randomized participants with available outcome data. Participants missing two or more follow-up time points were excluded. Descriptive statistics are presented as means and standard deviations or as frequencies and percentages as appropriate. Post-hoc subgroup analyses were performed with objective subgroup classification criteria to assess patients with severe (defined as worse than the median) baseline scores of back and leg pain, EQ5D, ODI, and initial scar score. Exploratory subgroup analyses were performed based on the following hypotheses: 1) patients with severe baseline functional impairment may have greater inflammatory burden as a substrate for photobiomodulation’s anti-inflammatory effects; 2) patients with severe initial scar scores may represent those with impaired early healing capacity in whom photobiomodulation may better facilitate perceivable improvement. Linear mixed-effects analyses were applied to all primary and secondary outcomes including scar score, VAS back pain, VAS leg pain, EQ5D, and ODI for both the overall cohort and subgroup analyses using Python (Python Software Foundation). Group and time were treated as fixed effects, and subject variability was controlled as random effect. Mixed-effects models used all available repeated measures per participant. Post-hoc pairwise comparisons were performed in Prism 10 (GraphPad, San Diego, USA) using Sidak’s correction, with families defined by time points and groups, following a mixed-effects model to evaluate treatment effects. A nonlinear mixed-effects model with an asymptotic exponential function was developed to predict the healing trajectories over time using R version 4.0.3 (R Foundation for Statistical Computing). Model uncertainty was quantified using a parametric bootstrap with 1000 simulations of fixed-effect parameters based on model estimates. Descriptive values are expressed in mean ± SD. A p < 0.05 was considered significant.

## RESULTS

A total of 46 patients with lumbar decompression surgery were enrolled, and 25 patients met inclusion criteria for this study. Among the 21 excluded, 5 converted from original intended surgical procedure (i.e. minimally invasive to open surgery), 11 did not provide any follow-up data, and 5 exceeded the prespecified missing-data threshold. Among the 25 included patients, 13 were randomized to the active photobiomodulation brace group, and 12 to the placebo brace group (**Figure 1**). One patient in the placebo brace group did not report VAS back pain scores at 6-weeks. Two patients in the placebo brace group and three patients in the active photobiomodulation brace group did not submit scar photographs at 2 weeks, and three patients in the active photobiomodulation brace group did not submit scar photographs at 6 weeks. **Table 1** summarized patient demographics and comorbidities. Surgical characteristics and wound closure–related variables were comparable between groups, with no significant differences observed (**Supplemental Table 1**).

**Table 1.** Demographics and surgical location of trial subjects by treatment group.

|  | Placebo (N=12) | Treatment (N=13) |
| --- | --- | --- |
| Mean age±SD, years (range) | 55±18 (28-81) | 59±12 (38-75) |
| Male/female | 9 /3 | 9 /13 |
| Comorbidities: |  |  |
| BMI > 30 | 2 | 1 |
| Hypertension | 5 | 8 |
| Hyperlipidemia | 7 | 5 |
| Coronary artery disease | 2 | 0 |
| Peripheral vascular disease | 0 | 0 |
| Anxiety | 4 | 4 |
| Depression | 5 | 0 |
| Diabetes mellitus | 1 | 1 |
| History of cancer | 2 | 5 |
| Thyroid dysfunction | 3 | 2 |
| Hematological disorders (bleeding/clotting) | 0 | 1 |
| Smoker (current/former) | 0/ 4 | 0/3 |
| Unemployed/retired | 1 /5 | 1/0 |
| Employed | 5 | 8 |
| Self-employed | 0 | 3 |
| Student | 1 | 1 |
| Level operated: |  |  |
| L2-3 | 1 | 0 |
| L3-4 | 3 | 3 |
| L4-5 | 4 | 9 |
| L5-S | 4 | 1 |
| Primary or Revision surgery | 1 | 0 |
| Complications (SSI, readmission) | 0 | 0 |

Abbreviations: SD=standard deviation; BMI=body mass index; SSI=surgical site infection; N=number of patients.

There were no adverse events associated with the use of photobiomodulation treatment and no surgical complications including surgical site infection or readmission for any reason during the follow up period for both groups.

Scars were evaluated using the Stony Brook Scar Evaluation Scale, which includes five domains (width, height, color, suture, and appearance), each scored as 0 or 1, with higher scores indicating more favorable healing. Figure 2 shows representative scars from one patient in the treatment group (**Figure 2C**) and one in the placebo group (**Figure 2D**). For treated patients, total scores improved from 2 at week 2 (width: 0, height: 1, color: 0, suture: 0, appearance: 1) to 4 at week 12 (width: 1, height: 1, color: 0, suture: 1, appearance: 1). For patients from the placebo group, scar scores were 3 at week 2 (width: 1, height: 0, color: 0, suture: 1, appearance: 1) and remained unchanged at week 12 (width: 0, height: 1, color: 0, suture: 1, appearance: 1).

**Table 2** shows outcome scores at baseline and weeks 2, 4, 6, 8, and 12. There were no significant baseline differences in back pain, leg pain, EQ5D, and ODI (p=0.88, 0.88, 0.88, 0.43, respectively), indicating appropriate randomization.

**Table 2.** Scores at baseline and postoperative weeks 2, 4, 6, 8, and 12.

|  | **Treatment (N=13)**<br>**Mean Score ± SD** | **Placebo (N=12)**<br>**Mean Score ± SD** |
| --- | --- | --- |
| ***Visual Analog Scale (VAS) – Back Pain Score*** |  |  |
| Baseline | 5.92 ± 2.84 | 5.25 ± 2.30 |
| 2 weeks post-op | 3.46 ± 1.85 | 2.42 ± 1.62 |
| 4 weeks post-op | 2.46 ± 1.90 | 2.25 ± 1.86 |
| 6 weeks post-op | 3.00 ± 2.32 | 2.00 ± 1.65 |
| 8 weeks post-op | 2.08 ± 1.38 | 1.83 ± 1.40 |
| 12 weeks post-op | 1.23 ± 1.30 | 1.50 ± 1.51 |
| ***Visual Analog Scale (VAS) – Leg Pain Score*** |  |  |
| Baseline | 5.54 ± 3.07 | 5.75 ± 2.18 |
| 2 weeks post-op | 2.46 ± 2.37 | 2.08 ± 1.62 |
| 4 weeks post-op | 1.69 ± 1.49 | 2.92 ± 2.02 |
| 6 weeks post-op | 2.46 ± 2.44 | 2.25 ± 2.01 |
| 8 weeks post-op | 1.85 ± 1.57 | 1.83 ± 1.64 |
| 12 weeks post-op | 1.08 ± 2.22 | 1.42 ± 1.31 |
| ***EuroQol (EQ) 5D*** |  |  |
| Baseline | 51.69 ± 26.16 | 50.25 ± 21.80 |
| 2 weeks post-op | 66.00 ± 18.58 | 61.50 ± 23.99 |
| 4 weeks post-op | 71.77 ± 18.09 | 65.42 ± 19.87 |
| 6 weeks post-op | 72.85 ± 21.35 | 65.92 ± 18.55 |
| 8 weeks post-op | 74.08 ± 19.36 | 74.17 ± 18.23 |
| 12 weeks post-op | 83.77 ± 14.24 | 77.33 ± 19.39 |
| ***Oswestry Disability Index (ODI)*** |  |  |
| Baseline | 0.52 ± 0.19 | 0.40 ± 0.15 |
| 2 weeks post-op | 0.40 ± 0.21 | 0.35 ± 0.10 |
| 4 weeks post-op | 0.32 ± 0.18 | 0.30 ± 0.13 |
| 6 weeks post-op | 0.29 ± 0.19 | 0.26 ± 0.15 |
| 8 weeks post-op | 0.24 ± 0.14 | 0.19 ± 0.12 |
| 12 weeks post-op | 0.17 ± 0.14 | 0.17 ± 0.13 |
| ***Stony Brook Scar Score*** |  |  |
| 2 weeks post-op | 2.09 ± 1.04 | 2.78 ± 0.97 |
| 4 weeks post-op | 3.69 ± 0.75 | 3.25 ± 1.14 |
| 6 weeks post-op | 4.15 ± 0.55 | 2.89 ± 0.93 |
| 8 weeks post-op | 4.15 ± 0.55 | 3.33 ± 0.89 |
| 12 weeks post-op | 4.23 ± 1.01 | 3.33 ± 0.65 |

Abbreviations: SD=standard deviation; N=number of patients.

Linear mixed-effects analysis showed a three-fold faster healing rate in the treatment group compared with placebo (k_treatment = 0.44, k_placebo=0.15, p=0.029) (**Figure 3A**). Post-hoc pairwise comparisons demonstrated significantly higher scar scores in the treatment group at 6, 8, and 12 weeks after surgery (p=0.0028, 0.020, and 0.011, respectively) (**Figure 3B**). Within-group analysis showed scar improvement by 2 weeks in the treatment group (p<0.0001), whereas no significant change was observed in the placebo group. Linear mixed-effects analyses of the secondary outcomes did not find significant differences between treatment and placebo groups in EQ5D, back and leg pain, and ODI (**Supplementary Figure 1**).

**Figure 3.**
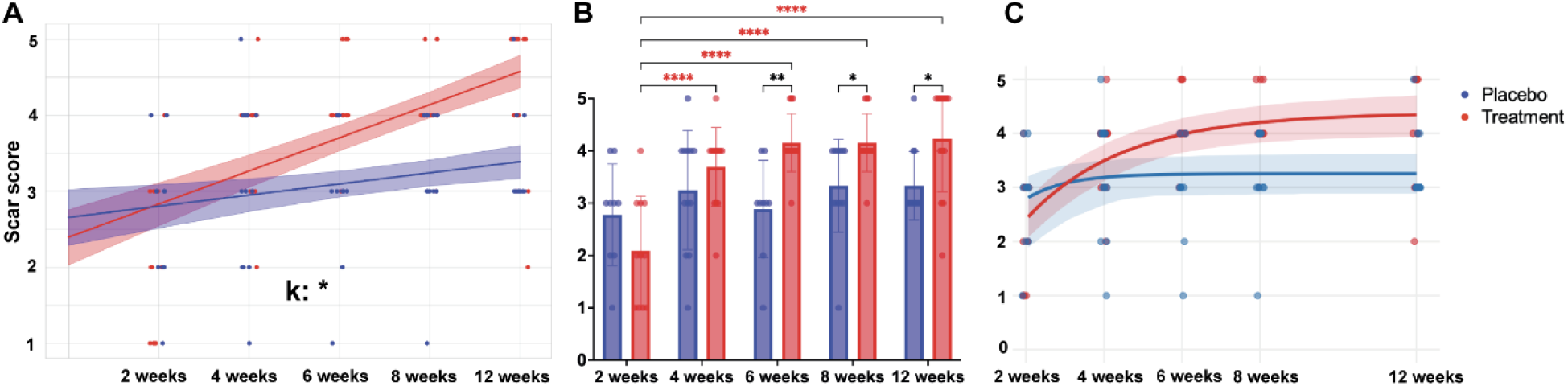
Treatment group showed significantly greater scar improvement compared to the placebo group. **A:** Linear mixed-effects analysis of scar scores over time. **B:** Post-hoc pairwise comparisons between groups at each time point. **C:** Predicated scar trajectories using a nonlinear mixed-effects model. Blue: placebo group. Red: treatment group. *: p < 0.05, **: p < 0.01, ***: p < 0.001, ****: p < 0.0001.

A nonlinear mixed-effects model using an asymptotic exponential function was developed to characterize biological scar healing trajectories. This model included scar score plateau and rate constant as fixed effects and subject as a random effect (**Figure 3C**). The model demonstrated moderate explanatory power with a marginal R^2^ of 0.37 (fixed effects only) and a conditional R^2^ of 0.53 (fixed and random effects), which indicates both treatment and individual variability play a role in the scar healing dynamics. In the placebo group, the estimated plateau scar score was 3.26, and the rate constant (k) was 0.97, indicating a relatively fast but limited healing response. On the other hand, treatment significantly increased the plateau score by 1.13 points (SE=0.29, p=0.0002), corresponding to a 4.12-fold accumulative improvement in final scar healing, although it takes longer to achieve the plateau (Δk = –0.57, SE=0.28, p=0.046). These findings suggest that without treatment, patients reached a plateau faster but at a limited healing capacity.

Subgroup analyses were performed to assess whether severe (defined as worse than the median) baseline scores of back and leg pain, EQ5D, ODI, and initial scar score were associated with differences in improvement. Among patients with severe preoperative baseline ODI, treatment group had 1.89-fold faster improvement in back pain score compared to placebo (k_treatment = -1.0, k_placebo = -0.53, p=0.025). Post-hoc comparisons showed that, while there was no significant between-group difference at individual timepoints, the treatment group demonstrated continuous significant back pain improvement over the 12-week period, whereas the placebo group improved initially at 2 weeks but plateaued thereafter (**Figure 4A, 4B**). This subgroup also demonstrated treatment effects on ODI, with laser brace group having 1.80-fold faster improvement in ODI score (k_treatment = -0.09, k_placebo = 0.05, p = 0.025); and similarly, only treatment group exhibited continued ODI improvement over time (**Figure 4C, 4D**). Treatment also resulted in borderline significant improvement in scar scores compared to placebo (k_treatment = 0.63, k_placebo = 0.24, p=0.055), corresponding to a 2.6-fold faster wound healing in the treatment group. Post-hoc pairwise comparisons showed significantly better scores in the treatment group at week 6, 8, and 12 compared to the placebo group (p = 0.0062, 0.010, 0.042, respectively) (**Figure 4E, 4F**).

**Figure 4.**
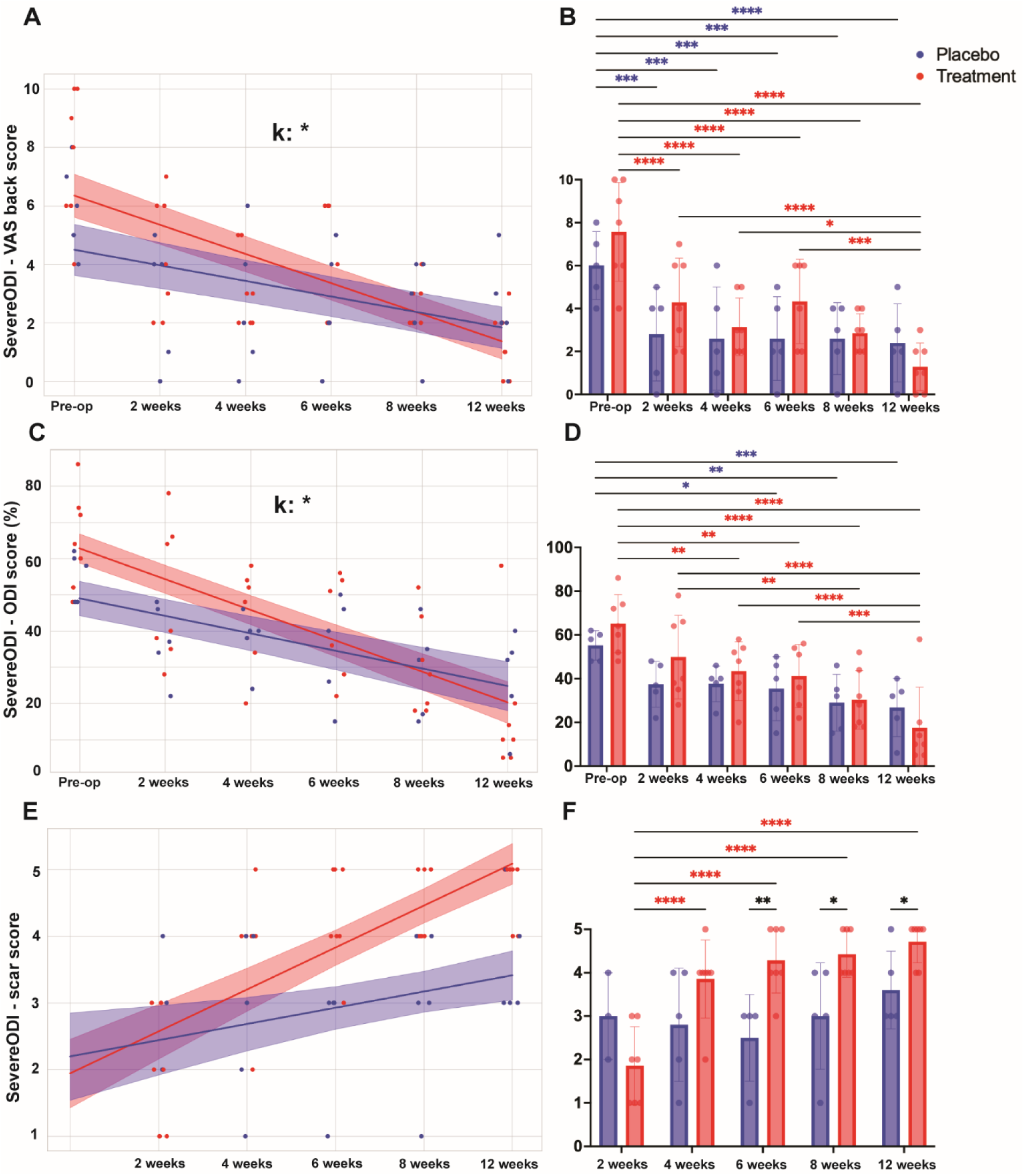
Subgroup analyses of patients with severe pre-op baseline ODI scores (above median). **A, B:** VAS back pain outcomes. **C, D:** ODI scores. **E, F:** Scar outcomes. Blue: placebo group. Red: treatment group. *: p < 0.05, **: p < 0.01, ***: p < 0.001, ****: p < 0.0001.

Among patients who had severe initial scar grades at two weeks after surgery, treatment produced significantly better scars at 6 weeks (p = 0.0034), and linear mixed-effects analysis showed a faster healing rate, although this did not reach significance (k_treatment = 0.69, k_placebo = 0.47, p=0.23) (**Figure 5A, 5B**). Post-hoc comparisons showed that scar improvement occurred by 2 weeks in the treatment group, whereas the placebo group did not show significant improvement until 6 weeks (**Figure 5B**). For EQ5D, the treatment group showed borderline greater improvement than placebo (k_treatment = 6.8, k_placebo = 3.5, p=0.052), corresponding to a 1.94- fold improvement (**Figure 5C**). Additionally, patients who received laser treatment also exhibited significantly improved EQ5D from baseline at 2 weeks, whereas the placebo group had no significant improvement over time (**Figure 5D**). The other subgroup outcome analyses did not demonstrate significant differences between treatment and placebo groups.

**Figure 5.**
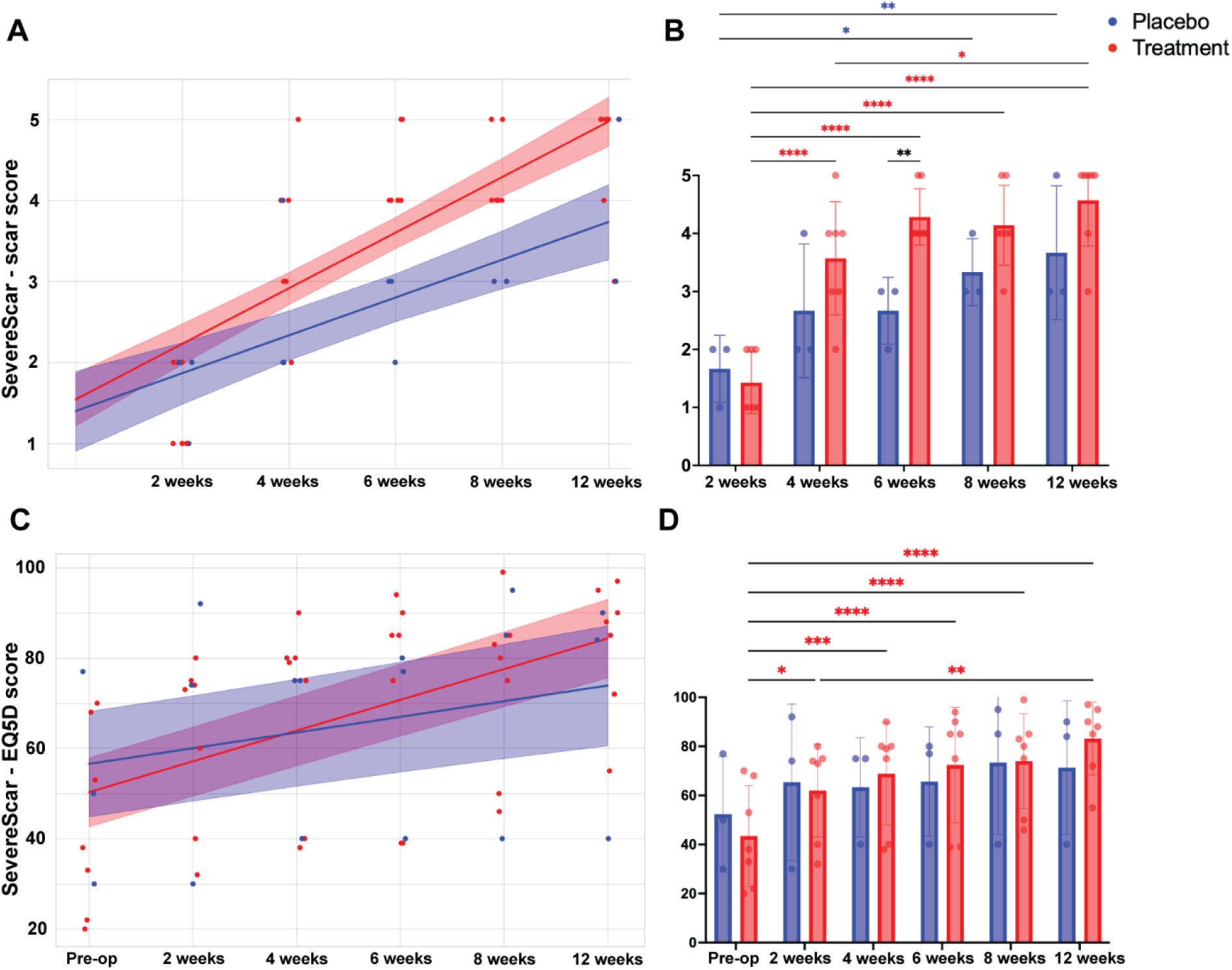
Subgroup analyses of patients with severe initial scar scores at post-op week 2. **A, B:** Scar outcomes. **C, D:** EQ5D outcomes. Blue: placebo group. Red: treatment group. *: p < 0.05, **: p < 0.01, ***: p < 0.001, ****: p < 0.0001.

## DISCUSSION

### Wound healing outcomes

This double-blinded, placebo-controlled study explored the effectiveness of laser therapy in enhancing the healing process for patients undergoing lumbar decompression surgery. Patients receiving photobiomodulation had significant improvement in scar healing at 6, 8, and 12 weeks, with early benefits detectable as soon as two weeks after initial scar assessment. No adverse effects were observed.

Exponential nonlinear mixed-effects model revealed that laser treatment significantly improved the final scar outcome, as evidenced by a higher scar score plateau. In contrast, the placebo group exhibited a lower asymptote and higher rate constant, suggesting that while natural wound healing progresses more rapidly towards a plateau, the final scar outcome was less favorable. The slower predicted healing rate of treatment group may reflect photobiomodulation- induced tissue regeneration process, such as collagen remodeling and angiogenesis, which may transiently delay the visible scar healing but ultimately leads to a better structural and cosmetic outcome.

### Functional outcomes and subgroup analyses

Subgroup analyses demonstrate that patients with severe preoperative disability experienced greater benefits from photobiomodulation, including faster reduction in back pain, faster functional recovery, and greater scar improvement, suggesting photobiomodulation may be particularly beneficial in patients with higher baseline disease burden.

Notably, the significant improvement in pain and functionality was only observed in subgroup but not the overall group analyses. There are several possible explanations. First, photobiomodulation predominately acts on inflammatory pathways, while pain perception can be influenced by multiple factors, including baseline pain severity and duration, disability, sensitivity, and age ^19^, which also helps explain why the pain reduction becomes significant in patients with more severe baseline disability. Second, the questionnaires for patient-reported pain do not distinguish deeper tissue and radicular pain versus wound pain. Third, our laser braces used only one dose and wavelength, which may not provide adequate penetration for deeper tissues ^6^. Furthermore, the surgical intervention itself could contribute to a substantial decrease in pain in both groups, confounding the effects of laser treatment. Fourth, the postoperative environment, including analgesia, activity restriction, and surgical inflammation, could also confound the perception of back pain. Reduction in post-surgical inflammation, rather than a direct analgesic effect on chronic back pain, may be the more mechanistically accurate interpretation of the observed pain benefits in this cohort. A non-operative patient population or a surgical cohort without pre-existing axial back pain would provide a cleaner test of photobiomodulation’s pain-modulating effects and is an important direction for future research.

### Mechanism of photobiomodulation

Photobiomodulation has emerged as a promising tool in the past two decades to facilitate pain management and wound healing ^4, 20, 21^. Photobiomodulation works by emitting low-intensity light, usually in the red or near-infrared range, to stimulate photoreceptors on cellular and mitochondrial membranes. Photobiomodulation enhances mitochondrial respiration ATP production and reduces production of pro-inflammatory cytokines ^22^, and increases re-epithelialization ^9, 23, 24^.

An important practical consideration is the depth of tissue penetration, which is directly governed by wavelength. Red light at 650 nm, as used in the current device, penetrates approximately 5-10 mm into tissue – sufficient to act on dermal and superficial subcutaneous structures but unlikely to reach the deeper paraspinal musculature, epidural space, or neural elements that contribute to axial back pain and radicular symptoms. Near-infrared wavelengths (800-1100 nm) achieve penetration depths of 2-3 cm or greater and are better suited for targeting deeper tissues. Cytochrome c oxidase absorbs red to near-infrared light within the 600-1000 nm wavelength range, which is thought to modulate nociceptive signaling pathways. Improvements in functional capacity, often, though not exclusively, related to reductions in pain, may occur concurrently. This wavelength-penetration relationship is a critical consideration in interpreting the differential effects observed in this study: robust improvements in superficial wound healing with more modest and inconsistent effects on pain and functional outcomes are mechanistically consistent with the depth limitations of 650 nm light. Future devices targeting deeper spinal tissue should incorporate near-infrared wavelengths and higher output power.

### Photobiomodulation in current literature

Several prior randomized controlled trials have established the efficacy of photobiomodulation in postoperative settings across surgical specialties. In patients undergoing total hip arthroplasty, direct laser therapy applied to the surgical scar produced significantly lower VAS pain scores and lower inflammatory cytokines compared to placebo ^25^. Another study on postoperative pain of breast augmentation surgery showed that laser diode treatment immediately before and after surgery was associated with significantly reduced postoperative pain and less use of pain medications ^26^. A study on bariatric surgery wound revealed that patients in the laser group had lower wound temperature, lower erythrocyte sedimentation rate, reduced seroma, and improved cicatrization ^27^.

Improvement in wound healing is associated with multiple benefits. First, it decreases the risk of surgical site infection, a common incisional complication that accounts for 17-22% of healthcare-related infections, 9.7 days longer hospital stay, and annual cost of $3.3 billion ^31–34^. For spine surgeries, wound infection can further lead to hardware failure requiring reoperations. Improved wound healing also promotes early mobilization and rehabilitation and contributes to better functional outcomes and patient satisfaction. Furthermore, our subgroup analyses showed that among the patients with severe scars at the initial visit, laser treatment still exhibited superior effects on promoting scar healing compared to placebo, suggesting that the therapeutic benefit of photobiomodulation is not limited by the initial scar condition. This is particularly relevant to patients with comorbidities such as obesity, diabetes, immunosuppression, and cancers that predispose them to a poor initial wound ^35–37^. In diabetic wound models, for instance, photobiomodulation accelerates healing by increasing expression of vascular endothelial growth factor (VEGF), promoting capillary formation, and modulating oxidative stress and inflammatory signaling ^23^. In spinal metastasis, which often requires chemoradiation following surgery ^39–41^, wound healing is a major concern. One study found that patients with shorter interval between surgery and postoperative chemoradiation therapies have a higher risk of wound infection or delayed wound healing ^45^. Photobiomodulation may play a role in this population, allowing timely start of chemoradiation for disease control while reducing the risk of wound complications.

In the context of pain management, Konstantinovic et al. showed that in patients with acute low back pain with radiculopathy, laser therapy in adjunct to non-steroidal anti-inflammatory drugs resulted in significantly more reduction in pain and improvement in mobility and quality of life, compared to medication alone ^29^. A study on patients with chronic failed back surgery syndrome found that patients receiving 3 weeks of laser therapy had a 51% and 27% reduction in Numeric Rating Scale (NRS) pain score and Oswestry Disability Index, respectively, and such improvement was maintained at 6 months after treatment ^30^.

### Limitations and future directions

Several limitations should be considered. First, the study was limited to a 12-week follow-up period, which may not reflect long-term scar maturation or functional recovery. It is unclear whether there is continued benefit to skin healing beyond 12 weeks. Conversely, intervention-specific analgesic benefit may have become significant after the 12-week point, which our study did not capture. Second, the laser therapy was self-administered, so if participants did not strictly adhere to the protocol, differences in individual compliance could confound the results. Third, this study has a relatively small sample size, particularly the subgroup analyses, which raises the possibility of underpowered effect size estimates. The relatively small sample size was a consequence of our intention to maximize statistical rigor by excluding patients who had incomplete data. Given the limited sample size, a relatively small number of participant withdrawals resulted in a proportionally large exclusion rate, which may also be indicative of real-world challenges related to patient retention. Accordingly, our analysis more closely aligns with a per-protocol approach rather than an intention-to-treat framework. Although this methodology ensures a complete dataset for analysis, it may introduce bias and restrict the generalizability of the findings. Nevertheless, it may also offer insight into treatment effects under conditions that more closely reflect typical patterns of patient adherence.

While the subgroup analyses were defined to investigate patients with severe baseline functional status and pain, these findings may be subject to inflated effect estimates due to small subgroup samples. Further, there are other potential confounders, including surgical complexity, postoperative rehabilitation, inter-surgeon variability in wound closure technique despite institutional standardization, which could not be fully controlled given the small sample size. Therefore, these subgroup results should be interpreted as exploratory only. Additionally, the relatively high post-enrollment exclusion rate due to missing data and loss of follow-up may introduce selection bias. Finally, this study enrolled only patients undergoing minimally invasive lumbar decompression, and findings may not generalize to open surgical approaches or other spinal procedures.

The current pilot study lays the groundwork for several important future investigations. First, a larger, confirmatory randomized controlled trial is needed with a formal a priori power calculation based on the effect sizes observed here. Such a trial should extend follow-up beyond 12 weeks to assess durability of wound healing and potential delayed pain and functional benefits. Second, because the present cohort consisted of minimally invasive procedures with inherently low wound complication rates, the observed treatment effects may represent conservative estimates of the true clinical impact of photobiomodulation. Future studies may explore photobiomodulation in higher-risk surgical populations, including open lumbar fusion, deformity correction, and revision surgery. Third, systematic evaluation of alternative wavelengths (particularly near-infrared, 800-1100 nm) and higher-output devices are warranted to test whether deeper tissue penetration translates into more robust improvements in pain and functional outcomes. Fourth, objective monitoring of device use and compliance should be incorporated to enable dose-response analysis and control for differential adherence.

## CONCLUSIONS

This double-blinded, placebo-controlled study found that photobiomodulation treatment after lumbar decompression surgery led to significantly better wound healing at 6, 8, and 12 weeks post-surgery, with a 4.12-fold accumulative improvement in final scar scores compared to the placebo group. Subgroup analyses further revealed that among patients with severe preoperative disability, treatment provided a 1.89-fold faster improvement in back pain and a 1.80-fold faster improvement in ODI scores. There were no adverse events associated with this treatment. These therapeutic effects are likely mediated by photobiomodulation-induced reduction of inflammation and enhancement of tissue repair. Together, this study suggests that photobiomodulation may be a promising adjunct therapy to promote postoperative recovery in patients undergoing spine surgeries.

## ACKNOWLEDGMENTS

This study was made possible by the generous support of the Chen Family. The content is solely the responsibility of the authors and does not necessarily reflect the official views of the Chen Family. We thank Dr. James C. Leiter for helpful discussions and guidance on statistical analysis. We also acknowledge the late Dr. Irene Say for her foundational contributions to the conception and original design of this study.

## DATA AVAILABILITY STATEMENT

Individual de-identified participant data, data dictionary, and statistical code are not publicly available but may be shared upon reasonable request with institutional approvals.

## CONFLICT OF INTEREST

The authors report no conflict of interest related to this study.

---

**Supplemental Table 1.** Comparison of wound closure-related variables between placebo and photobiomodulation groups, including procedure type, surgical approach, and level of training of the individual performing closure. No significant between-group differences were observed.

| **Surgical variable** | **Placebo (N=12)** | **Treatment (N=13)** | **P-value** |
| --- | --- | --- | --- |
| Single level decompression | 11 (91.6%) | 13 (100%) | 0.99 |
| Multiple level decompression | 1 (8.4%) | 0 (0%) | 0.99 |
| Education level of person closing | Fellow: 5<br>PGY-5: 0<br>PGY-3: 6<br>PGY 2: 1 | Fellow: 4<br>PGY-5: 1<br>PGY-3: 4<br>PGY 2: 4 | 0.36 |
| Tubular retractor use | 12 | 13 | 0.99 |
| Laminectomies | 8 | 12 | 0.16 |
| Hemilaminectomies | 4 | 1 | 0.32 |
| Post-op narcotic use | 10 | 11 | 0.99 |
| **Surgical levels** |  |  |  |
| L2-3 | 1 | 0 | 0.48 |
| L3-4 | 3 | 3 | 0.99 |
| L4-5 | 4 | 9 | 0.09 |
| L5-S1 | 5 | 1 | 0.09 |

**Supplementary Figure 1.**
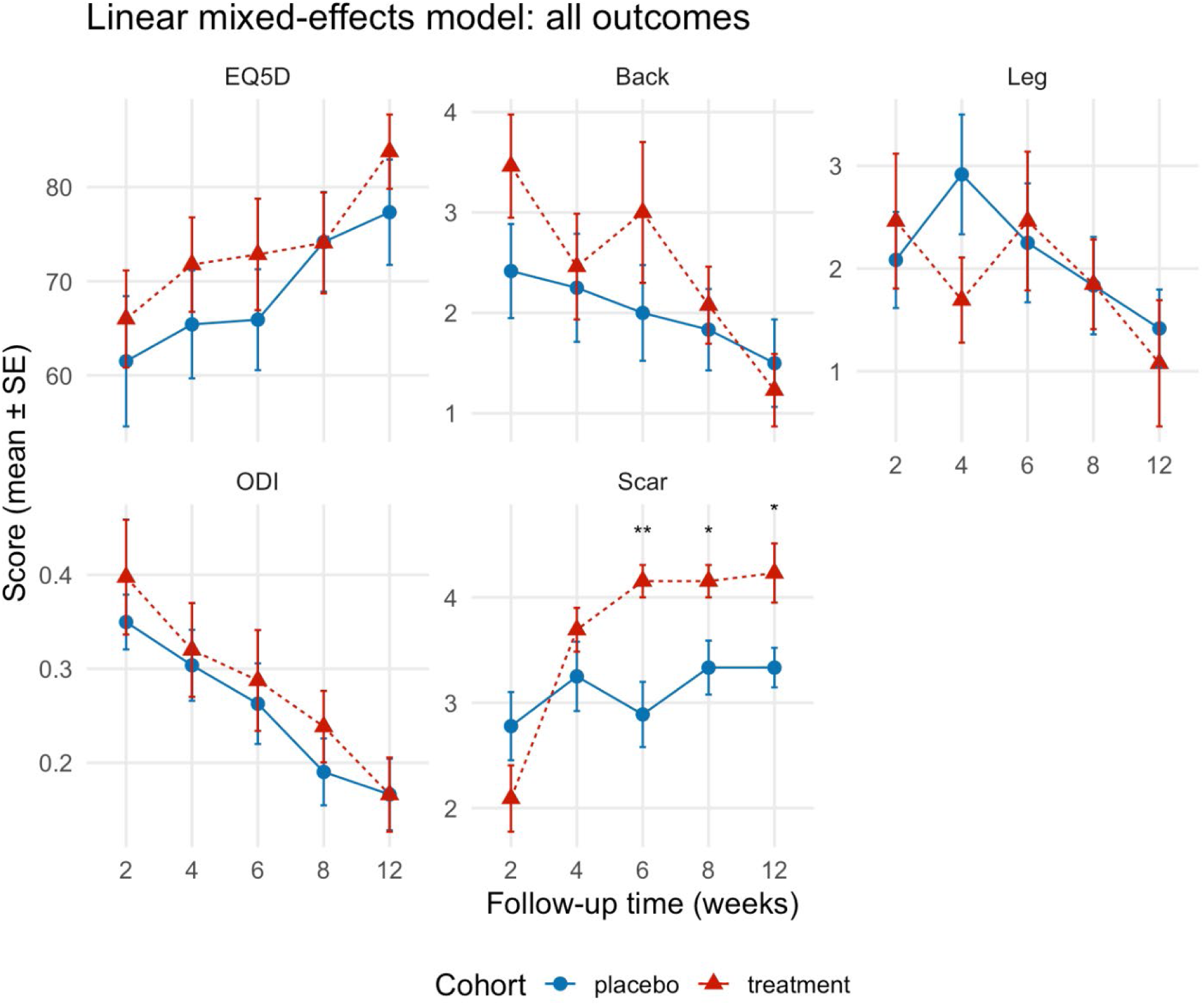
Linear mixed-effects analyses of secondary outcomes.

## Notes

### Competing Interest Statement

The authors have declared no competing interest.

### Clinical Trial

NCT06282770

### Author Declarations

IRB of University of California, Los Angeles gave ethical approval for this work (IRB#23-000444)

